# Student Cognitive Enhancement with Non-Prescribed Modafinil. Is it Cheating?

**DOI:** 10.1101/2024.03.01.24303594

**Authors:** Alexia Kesta, Philip M. Newton

**Affiliations:** Swansea University Medical School, Singleton Park, Swansea, Wales. SA2 8PP. United Kingdom

**Keywords:** modafinil, smart drugs, study aids, cheating, cognitive enhancers, neuroenhancment, academic integrity

## Abstract

Modafinil, a prescription-only drug, it is mainly used to treat narcolepsy and sleep disorders, but it is also used, without a prescription, as a cognitive enhancer by ∼10% of UK University students. Previous research has focused on the prevalence of, and motivations for, these behaviours. Here we focused specifically on determining whether students view this behaviour as cheating. We used a scenario-based approach to quantify, and qualitatively understand, student views on this topic. Most students did not view this behaviour as cheating, in part due to similarities with freely available stimulants such as caffeine, and a view that cognitive enhancement does not confer new knowledge or understanding. Although a minority of students did view it as cheating, they also expressed strong views, based in part on basic questions of fairness and access. Few students did not have a view either way. These views remained largely unchanged even when presented with considerations of other moderators of the ethics of cognitive enhancement with modafinil.

## Introduction

Neuroenhancement is broadly defined as the use of drugs or other interventions to “modify brain processes with the aim of enhancing memory, mood and attention in people who are not impaired by illness or disorder” (1). Cognitive enhancement falls under neuroenhancement and is a term normally used to describe the use of certain stimulant drugs, without a prescription, for the purposes of enhancing performance on cognitive tasks such as university assessments. Commonly cited cognitive enhancers include modafinil (Provigil®), methylphenidate (Ritalin®, Concerta®), and d-amphetamine (Adderall®) (2,3). Modafinil is a front-line treatment for narcolepsy, and is only available, in the UK, with a prescription (4). However, it is generally well tolerated and with a low risk of harm or abuse, and so it is often subject to less stringent regulation than other prescription stimulants. In the United Kingdom, for example, Modafinil is Schedule IV(II), meaning that *not* illegal to possess modafinil without a prescription, although it is illegal to supply it (5). Although modafinil is only currently approved for the treatment of narcolepsy, it is widely prescribed for ‘off label’ uses such as the sleepiness associated with conditions such as multiple sclerosis and depression, and a 2004 report from the manufacturer estimated that 90% of prescriptions were for such uses (6).

There is concern about the non-prescription use of prescription stimulants as cognitive enhancers (CEs) by university students, with media coverage portraying it as being extremely common (7). Estimates vary for the number of students who take these medicines as cognitive enhancers. A recent review of the use of these medicines by university student in the UK found that only 6.9% of students have used them, although this number was higher for modafinil (9.9 %) compared to methylphenidate (3.3%) or dexamphetamine (1.9%) (8).

The exact mechanism of action of modafinil is not fully understood, but it seems to act through effects on catecholamine transport, and actions on orexin neurons which increase the hypothalamic release of histamine resulting in wakefulness and alertness (9). *In vitro* studies have identified that modafinil inhibits the reuptake of dopamine (10). Modafinil has a long half-life, of 12-15 hours (11). Some common or very common side effects of modafinil include; anxiety, irregular heartbeat, headache, insomnia, nausea and dizziness (10), although these are partially due to placebo/expectancy effects (12). Modafinil is cheap and widely available: current estimates are that a single dose costs approximately 1.00 GBP (1.21 USD, 1.15 EUR) and it is easily available online via the black market (13).

It appears that modafinil is a modest cognitive enhancer, despite all the attention that has been paid to it. A 2019 meta-analysis, which included studies up to July 2016, demonstrated that modafinil has a small but significant effect (g = 0.1) across a range of different standardised tests of attention, executive function, processing speed and memory. There was no significant difference between the cognitive functions, but when analysed individually the effect on processing speed was the largest (g = 0.2) while the effects on attention and memory were not statistically significant.

The cognitive effects of modafinil overlap substantially with those of caffeine, and the effect of modafinil has been described as being similar to ‘a strong cup of coffee’ (5), and it seems reasonable to propose that use of caffeine fits the definition of neuroenhancement as given by Hall above. Similarly, the magnitude of modafinil’s cognitive enhancing effects appears to be similar to that of acute exercise (g = 0.1), although the effects of exercise appear to be on different cognitive processes, and perhaps masked by some negative effects of exercise on some cognitive tasks (14).

The use of smart drugs by university students creates an ethical question; is it cheating? Some researchers have argued that it is, and the most often expressed objections to the ethical use of CEs among academics and the general public are those relating to justice, safety, and coercion (15), rather than cheating. For example, healthy individuals who do not have any kind of disorder, but who take modafinil, give them an unfair advantage since they are able to study for longer with higher cognitive processing speed. This issue is reflected in the wider literature on public attitudes to cognitive enhancement, which are more accepting of the use of cognitive enhancers in situations where their use is ‘restorative’ rather than ‘enhancing’, i.e. fixing a perceived problem rather than enhancing the normal performance of an otherwise healthy individual (16). This issue is also reflected in the status of cognitive enhancers as ‘doping’ agents in competitive sports. Modafinil is listed by the World Anti Doping Agency (WADA) as a banned substance and some competitive events, such as video gaming and chess, test their participants for use of cognitive enhancers (17).

However, it remains unclear whether the specific use of cognitive enhancers by university students constitutes a form of academic misconduct. There are various ways in which the use of prescription medications by students might be a concern to universities; health concerns, legality and crime, as well as academic integrity. A policy analysis in the USA showed that most universities considered the unauthorised use cognitive enhancers by students to be an issue of drug misuse, but almost none considered cognitive enhancers as part of their academic integrity policy (18). A similar study conducted in the UK showed that cognitive enhancers were almost entirely absent from consideration by any policy, either concerning academic integrity or drug misuse, even though the unauthorised use of cognitive enhancers could reasonably be considered to fit in the definitions of both (19). A qualitative study of policymakers in Australia generated similar findings; the use of cognitive enhancers was a health issue, rather than an academic integrity issue (20).

On the other hand, the authorised use of stimulants, including modafinil, has a long history in the military, although there is a paucity of academic research on modafinil use in the military (21). Modafinil is used to counter the cognitive effects of sleep deprivation, rather than treat narcolepsy. There is also clearly a commercial interest in developing the use of modafinil for cognitive enhancement; in 2018 the energy drink manufacturer Red Bull filed a patent for the use of a modafinil analogue known as ‘CE-123’ to be used for improving motivation, cognitive functions, and ‘reference memory’ (22).

Thus the aim of this study was to determine whether university students viewed cognitive enhancement with modafinil as a form of cheating. We achieved this though the use of a scenario-based survey study. We captured an initial view on this, and then also evaluated student views on factors which have been reported to influence considerations of whether or not modafinil use is view as cheating, namely whether a modafinil-enhanced performance is an authentic representation of true performance, the likelihood of coercion into modafinil use, equality of access, and similarities to other enhancers such as caffeine (15). We then asked them again whether modafinil use should be considered as cheating.

## Methods

***Ethical Approval*** was obtained from Swansea University’s Medical School Research Ethics Sub-Committee (<u>SUMS RESC</u>) (ethics approval number: 2022-0125).

### Participants and Inclusion Criteria

Participants were recruited using the online participant pool www.Prolific.co. Inclusion criteria for participant were that they should be (1) university students in the UK, in at least in Year 2 of studies (to ensure they had some experience of undertaking assessments in Higher Education, but (3) not studying professional programmes (e.g. Medicine, Dentistry, Nursing, Law) due to the complications of the ‘Fitness to Practice’ principles, wherein students and graduates have to demonstrate a commitment to ethical standards which may influence their attitudes to cognitive enhancing drugs, and also places a duty of responsibility on them to act should they admit to it.

### Recruitment

The Prolific advert stated that participants were being sought for an anonymous survey on their views on the use of modafinil as a cognitive enhancer by UK university students. They were warned of the attention checks, and that they would be paid at the Prolific recommended ‘good’ rate of £9/hour. A pilot test of 10 participants was made on February 10 2023, to test the functionality of the Qualtrics/Prolific interface and to calibrate the duration of the study for payment purposes. No changes were needed following the pilot and so a further participants were recruited and completed the study on February 13 2023.

### Survey instrument

This was built in Qualtrics (*Qualtrics XM - Experience Management Software*, 2023). We used a forced response approach wherein participants had to answer a question before proceeding to the next question. The structure of the study instrument was as follows, and a copy of the instrument is available as supplementary material S1.

A Participant Information Form contained all necessary information about the study, including who is undertaking the research, what happens if they took part and any risks. There was also data protection and confidentiality information. Participants were informed that, by clicking ‘next’ to progress to the next page, they were providing consent.

Section one was designed to ensure that participants fully understood the effects of modafinil, including side effects, it’s legality, availability and cost. For each of those issues, participants were requested to read some given information and then, on the next page, to answer questions about what they had read. Participants were required to answer correctly to proceed to the next section. This also served as an attention check and quality control test for the data; participants were informed that if they did not answer correctly then they would not be paid.

### Section two. Participants were given the following scenario

*“A university student is worried that they are not going to pass their end-of-year exams. They have heard that modafinil, a drug prescribed for the sleep disorder narcolepsy, can also improve cognitive performance in people who do not have narcolepsy. The student does not have narcolepsy, but orders some modafinil online; it is not technically illegal to buy modafinil without a prescription, although it is illegal to sell it. The student takes the modafinil before the exam, and the modafinil makes them less tired, and improves their concentration, decision-making and planning”*.

Participants were then asked to rate their agreement with the following statements on a 5 point Likert scale: *“The student has cheated by taking modafinil”.* Participants were then asked to explain their response, using a free text box.

Participants were then asked two questions on issues which have been demonstrated to affect people’s perceptions of whether non-prescribed modafinil use is cheating (15). These were (a) whether or not the academic performance of a student who has taken modafinil can be considered authentic, (b) that by making modafinil use common, it might result in the coercing of others to take it. In both cases the original scenario was presented and then participants were asked the aforementioned question using a Likert scale.

Next it was explained that, according to the UK Charity ‘Drug Science’ the actions of modafinil have been linked to a very high dose of caffeine (5). After this information was given, participants had to answer a question on modafinil’s mechanism of action, again to check their understanding and serve as an attention check.

Participants were then asked if they had ever used modafinil, and then if so whether it was via prescription for a legitimate health condition.

The original scenario was then presented and participants were asked if they had changed their mind, and so again asked to rate their view on whether modafinil use was cheating

Finally participants were asked further information about why they had made their decision on whether or not modafinil use is cheating. First they were presented with four issues which have been known to affect people’s view on whether modafinil is cheating, and then asked to rank those from most important to least important. The issues were (a) comparison with caffeine, (b) authenticity of student performance when taking modafinil, (c) equality of access to modafinil, (d) likelihood of coercion. They were also given a fifth option of ‘other’, and then asked to explain their answer in a free text box.

Participants were then debriefed and paid.

### Data Analysis

Data were downloaded into a Microsoft Excel spreadsheet. Statistical analysis and figure creation was undertaken using Graphpad Prism v10 (Thousand Oaks, CA, USA). Likert scale responses were tested for agreement or disagreement by converting the responses to a 1-5 scale and then comparing the distribution of responses to the midpoint of the scale using a one-sample Wilcoxon signed rank test. Ranking data were analysed using a one-way repeated measures ANOVA for ranks (Friedman test), followed by *post hoc* Dunn’s multiple comparisons tests. Qualitative data were aggregated into a Microsoft Word document and then analysed using established six-step bottom-up thematic analysis (23). Step 1 was ‘familiarisation’, where the transcripts were repeatedly read, with notes made. Step 2 was ‘coding’, where common phrases (keywords) were highlighted and then grouped into codes. In Step 3, similar keywords were grouped into themes. In Step 4 all the themes were checked and re-examined to ensure that all the data provided were found and used in the analysis. In Step 5 the themes were given names and explanations, and in Step 6 they were incorporated into the manuscript. Steps 1-3 were undertaken by one author (XXX) and then Steps 4-6 undertaken through discussion and agreement between both authors.

## Results

### Is it cheating to use modafinil for cognitive enhancement

When participants were asked the first time whether the use of modafinil represented cheating, 72.5% selected either ‘somewhat disagree’ or ‘completely disagree’, while 21.5% selected ‘completely agree’ or ‘somewhat agree’. Only 6% were unsure. The distribution of responses was significantly different to the midpoint of the scale (W= -11881, p<0.0001). After participants had been presented with all the relevant content regarding the four influencing factors, they were asked this question again. The results were similar, with 74% selecting ‘somewhat disagree’ or ‘completely disagree’, and 17.5% selecting ‘completely agree’ or ‘somewhat agree’. This time 8.5% were unsure. Again the disagreement was significant, (W=-12125, p<0.0001).

Despite the similarity between the distributions in the before and after questions, there was some suggestion that the difference between them was significant. A large number (159/200) of the pairs were tied (i.e. the participants picked the same option in both before and after questions). A Wilcoxon signed rank test which included these (using Pratts method) returned a P value of 0.066 (W = -2047). However, when these ties were excluded the P value was significant (P = 0.036, W = -298), suggesting that those who had changed their response were even less likely to view the use of modafinil as cheating.

The results are shown in Fig 1.

**Fig 1.**
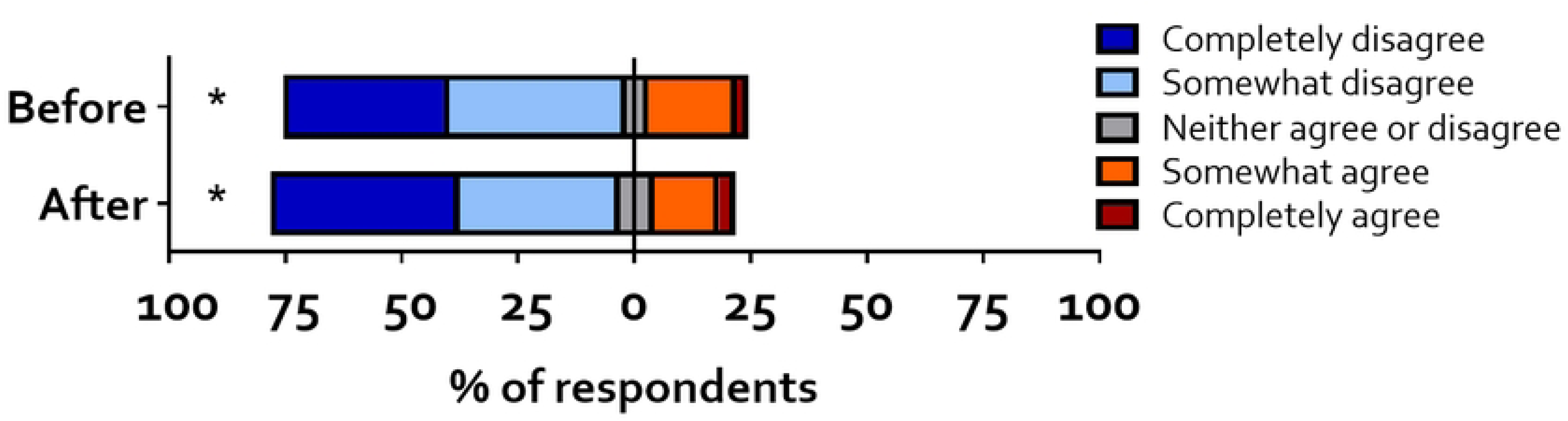
Is the student portrayed in the scenario cheating by using modafinil. Both distributions are significantly different to the midpoint of the scale, indicating significant disagreement with the statement that the student was cheating.

When asked if “the performance of the student in the scenario, who has taken modafinil, is not an authentic reflection of the student’s true academic performance”, the distribution of responses was significantly different (W= -5270 and P<0.0001). These results clearly suggest that participants showed significant disagreement with the given proposal, i.e. that the student’s performance was considered authentic when taking modafinil. Then, participants were asked to rate their agreement with the given statement “if modafinil use became common, those who do not want to use it, might feel forced to do so (coerced) by others”. Again, the distribution of responses was significantly different (W= 4971 and P<0.0001), although this time the participants agreed with the statement. These findings are shown in fig 2.

**Fig 2.**
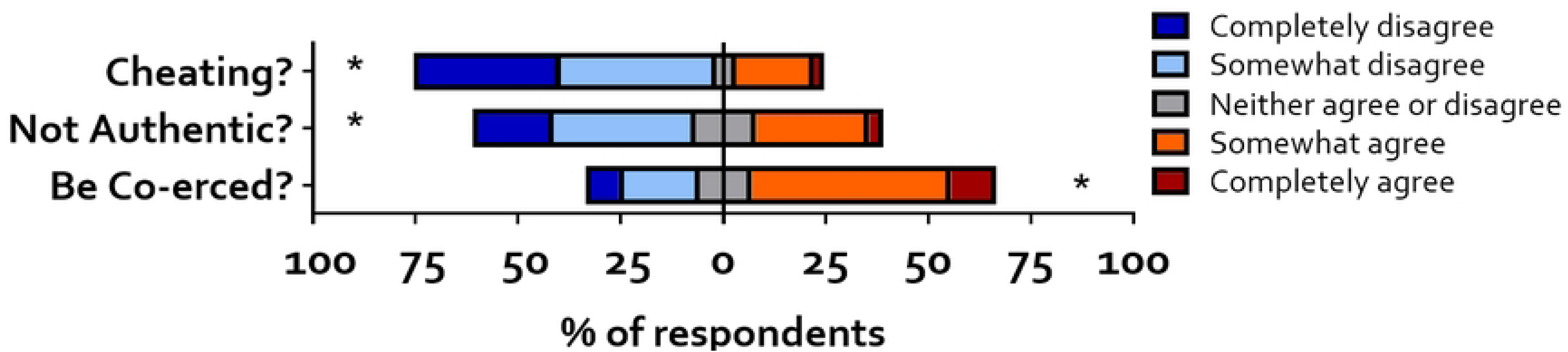
Distribution of answers to the Likert scale questions. Participants disagreed that modafinil use was cheating or represented an inauthentic portrayal of student performance, but they agreed that, if modafinil use became common, it might result in students feeling co-erced into using it.

When asked to rank the reasons why they had made their decision regarding the use of modafinil was considered cheating, a Friedman test indicated a significant difference between the five items. Post hoc Dunn’s multiple comparisons test showed that all items were significantly different to each other, except ‘Caffeine’ and ‘Authenticity’. These data can be seen in fig 3. The rankings were ordinal, but a mean was calculated for the purposes of visualisation (24). The mean rank for ‘caffeine’ was 2.26, for ‘authenticity’ 2.27, for ‘equality of access’ 2.72 and finally for ‘coercion’ 3.43.

**Fig 3.**
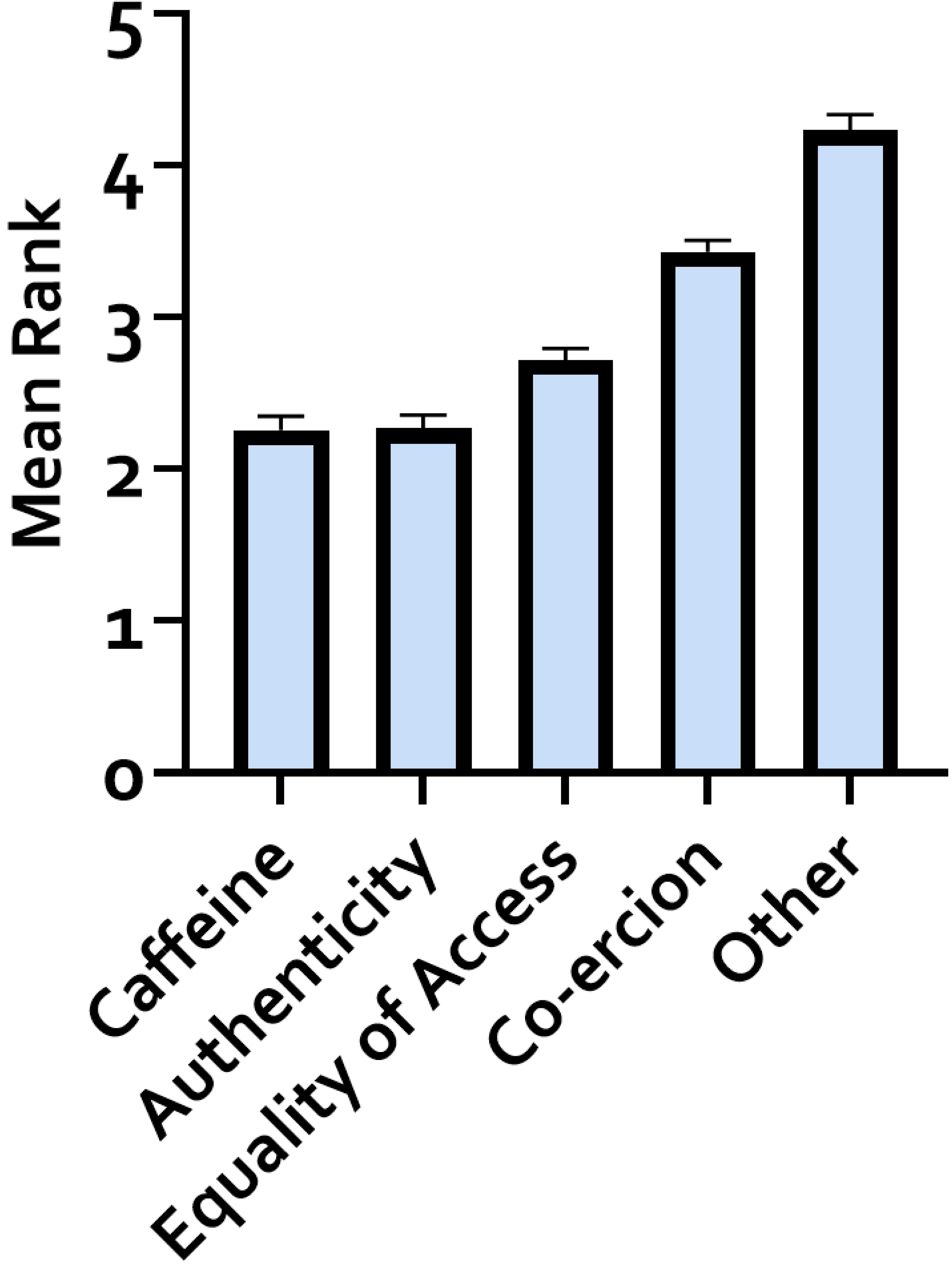
Reasons proposed for considering whether or not student use of modafinil in the scenario was cheating. Data are plotted as the mean ranking.

### Modafinil use

88% of participants reported that they had not used modafinil, while 12% (N=24) answered ‘Yes’. One (0.5%) participant had a prescription and so 11.5% did not obtain modafinil via prescription. Of the 24 who answered ‘yes’ to taking modafinil, 22 (92%) thought that it was not cheating when first asked and none of them changed their minds.

### Data quality

All who participated in the survey, answered the ‘knowledge’ questions correctly, and so no participants were excluded.

### Thematic Analysis

Participants were twice asked to explain their answers to the main question of whether modafinil is cheating, once at the beginning, and then again at the end after all the information had been presented. The results from the first question are shown in Table 1, along with representative quotes. The results and quotes from the second question are in Table 2. In both tables the keywords section are exact common words that participants used in their answers or comments, which were all mutual and categorised accordingly to their meaning. In Table 2 the themes essentially reflect the four items that participants were asked to rank, which are themselves the keywords, and so keywords/themes are collapsed into one column. It is important to be clear that the themes are not categorised into ‘positive’ and ‘negative’, meaning that some participants used the theme to support a view that taking modafinil is cheating whereas others used the same theme to argue the opposite.

**Table 1:** Themes arising from free text responses to the first asking of ‘is this cheating’. Table 1. Students were given a scenario where a student consumed modafinil before an exam, despite not having a prescription. Participants were asked whether or not they considered this to be cheating, and then asked why, in a free text box. These free text responses were analysed using thematic analysis.

| Theme | Keywords | Explanation | Example (phrase from participants) |
| --- | --- | --- | --- |
| **Attention** | Focus, concentration, awake, alertness, tiredness | The use of the drug affects cognition, improving the attention and awareness of the individual to help them study | "If they have a higher level of **concentration** usually, they would receive the same grade" "The role of the drug is to improve **concentration** and reduce **tiredness**" |
| **Intelligence** | Knowledge, study, performance, abilities | The use of the drug does not interact with the person's ability and how smart they are | "The drug does not help the student in any way that is unnatural or give them any extra **knowledge** to pass the exam" "It couldn't bring out any **knowledge** or **ability** which wasn't already there" |

Table 1. Students were given a scenario where a student consumed modafinil before an exam, despite not having a prescription.
|  |  |  |  |
| --- | --- | --- | --- |
| **Legality** | Rules, law, legal, illegal, regulations | The permission of students to take modafinil | "If it was cheating, there would be a **rule** against it"<br>"The law states it is not **illegal** to buy modafinil without a prescription and so the student has acted lawfully until this point" |
| **Consuming stimulants** | Coffee, tea, energy drinks, steroids | Stimulant compounds which can be used to improve alertness and performance | "They haven't technically cheated as it could be argued that a student **drinking energy** drinks before the exam is cheating" "It could be compared to a strong dose of **caffeine**" |
| **Solution** | Answers | Participants explained that by taking modafinil, it doesn't mean that you have the answers to the exam's questions, so it isn't cheating | "They haven't cheated by taking the drug as it does not make them have the **answers** to the exam questions" |
| **Well-being** | Health, unhealthy, risks, dangerous, diseases, side effects | The use of modafinil might affect the health of the person taking it | "The student is engaging in **unhealthy** behaviour that could end up in drug abuse or addiction" |
| **Factors** | Coercion,<br>advantage,<br>authenticity,<br>equality | Factors we gave participants<br>to help them decide if the use<br>of modafinil is cheating | "It *disadvantages* other students<br>who have not used the drug" "I<br>don't think that the student<br>taking this particular drug has<br>given them any significant<br>*advantages* compared to other<br>students" |

**Table 2:**
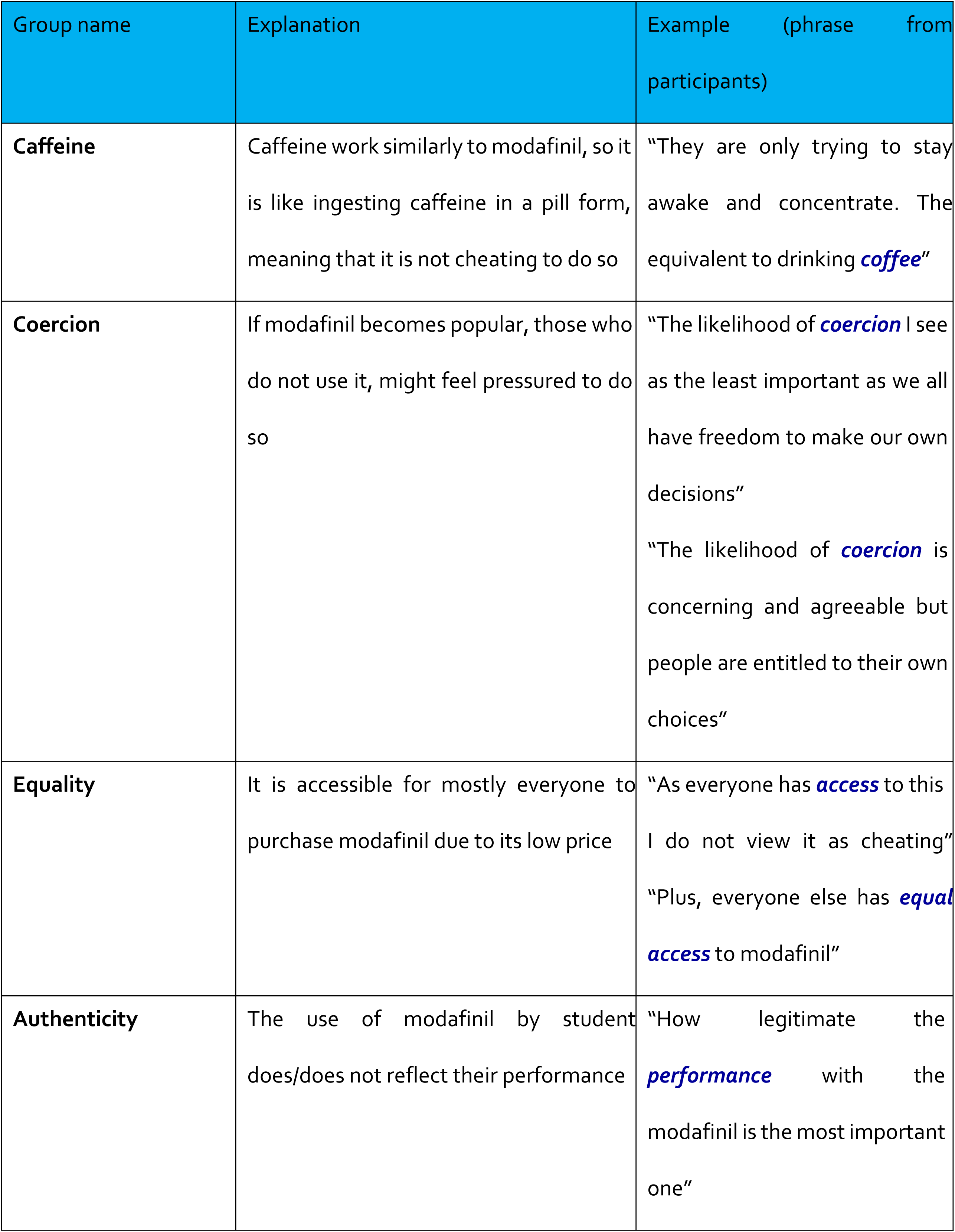
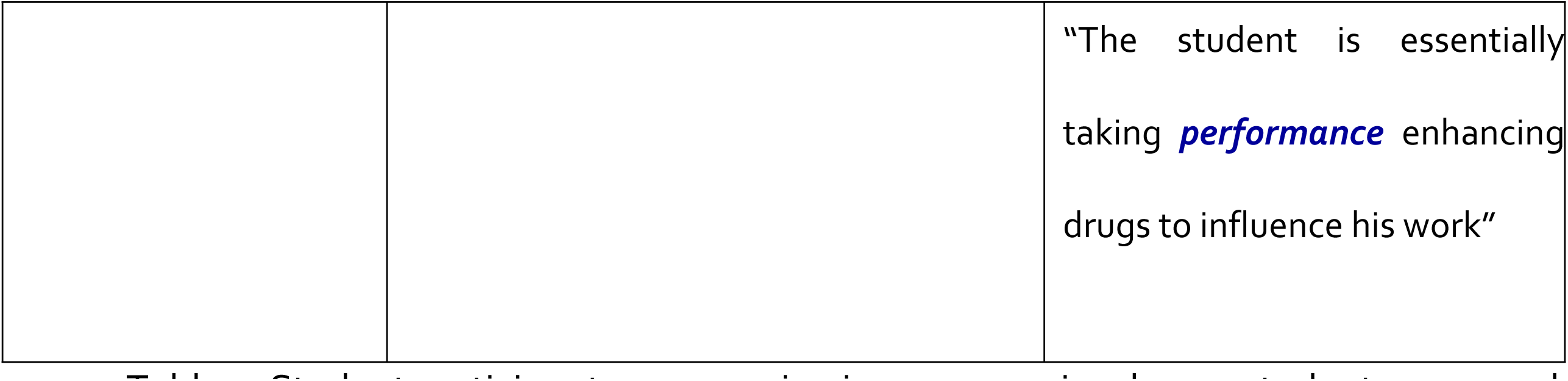
Themes arising from free text responses to the second asking of ‘is this cheating’. Table 2. Student participants were again given a scenario where a student consumed modafinil before an exam, despite not having a prescription. Prior to this they had been asked various questions about issues which have been demonstrated in the literature, to affect considerations of whether modafinil use is cheating. They were then asked if they had changed their mind on whether this was cheating, and then asked why, in a free text box. These free text responses were analysed using thematic analysis.

## Discussion

The present study explores the views of university students about whether the non-prescription use of modafinil by healthy non-sleep-deprived individuals, as a study aid, is considered cheating. A substantial and significant majority of students reported that they did not consider it cheating, and this view remained largely unchanged when they were given information about concepts that are commonly regarded, in the academic literature, to affect people’s views on the question. Common reasons for this finding were a comparison to legal non-prescription stimulants such as caffeine, and a related view that modafinil would not actually equip a student with new knowledge or skills. Although a minority of students viewed modafinil use as cheating, they expressed strong statements in support of their view, and only a small percentage of students (6-8%) were unsure. 11.5% of participants had used modafinil themselves, without a prescription, a figure in line with a recent review of UK university students (8). Student participants also disagreed that the performance of a student who had used modafinil was an inauthentic representation of the performance of that student. However they agreed that, if modafinil use became common, other students might be co-erced into taking modafinil even if they do not want to.

Our qualitative analysis gave us considerable insight into these findings. When participants were first asked to explain their views on whether the use of modafinil was cheating, our thematic analysis returned 7 different themes. The first dominant theme, ‘attention’, linked with the keywords ‘focus, concentration, awake, alertness and tiredness’ demonstrated that students had a good understanding of the effects of modafinil. The second theme ‘intelligence’, linked with the keywords ‘knowledge, study, performance and abilities’, along with the sixth theme of ‘solution’ showed that participants view that modafinil was *not* cheating was explained partly by a thought that the use of the drug does not interact with a person’s ability, intelligence or knowledge (although some took the opposite view). This is a logical view and is supported by the literature; although modafinil can produce a modest increase in cognitive functions such as processing speed, it does not ‘implant’ knowledge or skills and does not actually appear to significantly improve memory either (25,26).

It remains an open question as to whether modafinil works as a cognitive enhancer under academic assessment conditions, and if so, how it might work. For example, a student could, as represented in the study, take modafinil before a test. This would be expected to produce the improvement in cognitive functions described in the study scenario, but it is unclear whether this translates to improved grades on an assessment. To truly answer this question would require a randomised controlled trial of a modafinil vs placebo vs untreated control, with performance of university students measured on a range of different assignment formats and lengths. Alternately, or in addition, modafinil might simply extend the time available for students to study by increasing wakefulness. This is the common use in military circumstances (21). This seems unlikely to be an effective strategy long-term, as eventually the ‘sleep debt’ would need to be repaid, but in the days before, and during, an assessment it is possible that a student could benefit from the short-term use of modafinil. To truly answer this question would again require a much stronger and authentic experimental design than the laboratory studies currently available.

Specifically relating to cheating, it was clear from the qualitative findings that part of a consideration of cheating is simply whether the use of modafinil is allowed by the university, or by the law, perhaps independent of any broader moral or ethical concerns. This was reflected in the theme of ‘legality’, where participants focused on what the law states and how important the law and rules are to this subject. A characteristic quote was that “*the law states it is not **illegal** to buy modafinil without a prescription and so the student has acted lawfully until this point*”. This suggests that, for some participants, increasing the scheduling of modafinil, so that it becomes illegal to possess it without a prescription, would be sufficient to change their view of whether it is cheating and so might deter cheating. This might also explain why modafinil use is currently more common amongst university students than other cognitive enhancers such as methylphenidate and amphetamine since both those drugs are Schedule II in the UK, meaning it is currently illegal to possess them without a prescription. Conversely, the efforts by the company Red Bull to patent an analogue of modafinil for cognitive enhancement (22) suggest that, should they be successful and so a modafinil analogue becomes more widely and legally available, the use of it would be expected to increase considerably. This then relates to the fourth theme of the qualitative analysis, consuming stimulants, where participants drew parallels to legal stimulants such as coffee, tea, energy drinks, as well as to doping with steroids.

The final two qualitative themes both related more to a negative view of modafinil, in particular that there may be an imbalance of harm vs benefits, as in previous work (9) or even that this may lead to addiction or pathological long-term use, although this does not seem to be a significant risk currently (3). There was a related view that the perceived risk taken by students who engage in modafinil use in the short term creates a disadvantage for those students who are not engaged in modafinil use, reflecting earlier research (27).

These views did not really change, from a qualitative perspective, once all the information had been presented to the participants, and the qualitative analysis of the second set of data suggested that, if anything views had reinforced and coalesced around the similarities with caffeine and also the inability of modafinil to confer knowledge and skills to a student who did not already have them.

Our participants were drawn from the online participant pool Prolific, which appears to be a first for studies of this kind. Although the data quality checks were stringent, no participants had to be withdrawn, giving us confidence in the data and suggesting that this may be a valuable avenue for further research in this area. However there are some limitations to these findings; our participant pool was a modest size, although many other studies of cognitive enhancer use by UK university students use smaller samples than ours and still generate important findings (e.g. (28–31)).

Another potential limitation is that there are potentially other modifying factors which may affect whether a student views the use of modafinil as cheating. For example, modafinil is currently on the World Anti-Doping Agency list of banned substances and is therefore prohibited for competitors (32), including for situations such as chess where increased modafinil might reasonably be expected to provide an advantage. Indeed the evidence suggest that modafinil does produce a modest improvement in the performance of amateur chess players, equivalent to that of caffeine, and in both cases correlated with players taking longer time on each move (33), although even this improvement prompted some to argue that modafinil should *not* be banned from professional chess, in part due to the culture of suspicion that it might create (34) and older studies from 2012/13 indicate that very few players take it (0.2%), or believe that it can improve their performance (1.5%) (35).In addition, some older literature suggests that modafinil may be more effective at cognitive enhancement in individuals with a lower IQ, although still within the normal range (106 vs 115) (36) and this has been argued as an ethical reason to promote access to, and use of cognitive enhancers (37). Finally some very recent research suggests that people who use modafinil for cognitive enhancement score more highly on measures of inattention and procrastination, suggesting that they may have symptoms of Attention-Deficit Hyperactivity Disorder (ADHD), and be self-medicating, although odafinil is not currently recommended as a treatment for ADHD in the United Kingdom.

The complexity of these issues is an avenue for further study in this area, in both quantitative and qualitative domains, to fully understand the depth and breadth of the ethical issues surrounding the potential use of cognitive enhancers in education. This need has become more pressing given the recent efforts of Red Bull, to patent a modafinil analogue for cognitive enhancement (22).

There is also a need to undertake research with other stakeholder groups. The current legal position of modafinil is unusual amongst stimulant drugs; most of the banned stimulants on the WADA list are in schedule II according to UK drugs legislation and so their legal status re cognitive enhancement is very clear; it is illegal to have them without a prescription. The opposite is true for caffeine, which is legal and freely available, but it is equally clear what the legal and ethical position is. Modafinil is in something of a limbo as it is not illegal to possess without a prescription, yet it is not available over the counter. It would be helpful to have the views of policymakers, lawmakers, parents, academics and other stakeholders in order to consider whether the scheduling should be relaxed, and so students can take it without having to obtain it from the black market, and can be supported to do so, or else the scheduling should be increased, so making it harder to obtain and forcing students to obtain legitimate sources of help for any undiagnosed ADHD or other study struggles.

The current legal and regulatory position also creates challenges for academic policymakers. In the US, cognitive enhancement is prohibited by university drug and alcohol misuse policies, although it is largely absent from academic integrity policies (18). A similar view is help by academics in Australia (20). In the UK, university policies are almost completely silent on the issue, with cognitive enhancement not featuring in either drug misuse, or academic integrity policies (19). This is despite the fact that ∼10% of UK university students are using modafinil to support their studies, and the lack of any policy position from UK universities seems likely to add to any confusion facing students when deciding whether or not is it acceptable to use modafinil for cognitive enhancement.

University teachers have been surveyed regarding their views on cognitive enhancement, and this research has shown that academics tend to have a less favourable view of cognitive enhancement than students although this research has focused on the university teachers using the drugs themselves, and also did not specifically focus on whether or not the use of modafinil by students is considered cheating (27). Similarly A survey study of health professionals in New Zealand found that most did *not* agree that it was ethical for university students to use cognitive enhancers (38).

In summary, a sample of UK university students showed a large majority who believed that the use of modafinil for cognitive enhancement was not cheating, in large part due to perceived similarities with caffeine and the perception that modafinil use could not give a student knew knowledge or skills. However minority of students strongly believed that it is cheating, on the basis of basic questions over fairness, equality of access and harm.

## Data Availability

All data provided as supplementary material

## Acknowledgements.

N/A

## Supporting Information Captions

**Supporting Information S1. An anonymised version of the full survey.**

